# Negative regulation of *ACE2* by interferons *in vivo* and its genetic control

**DOI:** 10.1101/2020.04.26.20080408

**Authors:** M. Azim Ansari, Emanuele Marchi, Narayan Ramamurthy, Dominik Aschenbrenner, Carl-Philipp Hackstein, STOP-HCV consortium, ISARIC-4C Investigators, Shang-Kuan Lin, Rory Bowden, Eshita Sharma, Vincent Pedergnana, Suresh Venkateswaran, Subra Kugathasan, Angela Mo, Greg Gibson, Graham Cooke, John McLauchlan, Eleanor Barnes, John Kenneth Baillie, Sarah Teichmann, Alex Mentzer, John Todd, Julian Knight, Holm Uhlig, Paul Klenerman

**Author notes:** These authors contributed equally (M. Azim Ansari, Emanuele Marchi, Narayan Ramamurthy, Paul Klenerman). Correspondence to: M. Azim Ansari & Paul Klenerman, Peter Medawar Building for Pathogen Research, University of Oxford, Oxford OX1 3SY, or.

## Abstract

The SARS-CoV-2 pandemic has resulted in widespread morbidity and mortality globally. *ACE2* is a receptor for SARS-CoV-2 and differences in expression may affect susceptibility to COVID-19. Using HCV-infected liver tissue from 195 individuals, we discovered that among genes negatively correlated with *ACE2*, interferon signalling pathways were highly enriched and observed down-regulation of *ACE2* after interferon-alpha treatment. Negative correlation was also found in the gastrointestinal tract and in lung tissue from a murine model of SARS-CoV-1 infection suggesting conserved regulation of *ACE2* across tissue and species. Performing a genome-wide eQTL analysis, we discovered that polymorphisms in the interferon lambda (*IFNL*) region are associated with *ACE2* expression. Increased *ACE2* expression in the liver was also associated with age and presence of cirrhosis. Polymorphisms in the *IFNL* region may impact not only antiviral responses but also *ACE2* with potential consequences for clinical outcomes in distinct ethnic groups and with implications for therapeutic interventions.

## Introduction

Severe acute respiratory syndrome–coronavirus 2 (SARS-CoV-2) which results in coronavirus disease 2019 (COVID-19) is a positive-stranded RNA virus that causes a severe respiratory syndrome in a subset of infected individuals and has led to widespread global mortality.

Entry of coronaviruses into susceptible cells depends on the binding of the spike (S) protein to a specific cell-surface protein and subsequent S protein priming by cellular proteases. Similar to SARS-CoV-1, infection by SARS-CoV-2 employs *ACE2* as a receptor for cellular entry^1^. Viral entry also depends on *TMPRSS2* protease activity, and cathepsin B/L activity may be able to substitute for *TMPRSS2*^1^.

Epidemiological studies have indicated that the risk for serious disease and death from COVID-19 is higher in males, in older individuals and those with co-morbidities^2–4^ and it varies across ethnic groups^5^. Host genetic variation is important in determining susceptibility and disease outcome for many infectious diseases^6^ and it is likely to be important in determining SARS-COV-2 susceptibility and outcome. Polymorphisms in the host genome could drive differences in *ACE2* expression which may affect SARS-CoV-2 susceptibility and infection outcome. Thus a better understanding of *ACE2* expression, its regulatory mechanisms *in vivo* and its association with host genetics, especially during viral infection will provide insights on SARS-CoV-2 pathogenesis and help in repurposing antiviral drugs and development of vaccine strategies.

Host genetic variation could impact on differential immune responses to the infection^7–9^. The earliest immune defence mechanism activated upon virus invasion is the innate immune system^10^. Virus-induced signalling through innate immune receptors prompts extensive changes in gene expression which are highly effective in resisting and controlling viruses and subsequently prompt the activation of inflammatory and or antiviral immune effectors involved in virus clearance^11^. It has been shown that host genetics contributes to transcriptional heterogeneity in response to infections^7,8,12^, which underlies some of the differences in innate immune responses observed between individuals and the varying susceptibility to infection^13^. Therefore in the context of infectious diseases, it is of paramount importance to investigate infected tissue to observe infection-triggered immune response heterogeneity and to understand the role of host genetics.

To understand the heterogeneity of *ACE2* expression in the presence of RNA virus infection, to detect common regulatory mechanisms and assess host genetic factors influencing it, we used HCV-infected liver biopsies from 195 individuals and performed liver transcriptomics and host genome-wide genotyping. Investigating correlation of *ACE2* expression with other genes, we observed negative correlation of *ACE2* with interferon-stimulated genes (ISGs). Using an independent HCV infected liver biopsy dataset we replicated the same effect and observed down-regulation of *ACE2* after treatment with pegylated IFN-*α*. We observed the same pattern in gastrointestinal tract where inflammation driven ISGs were negatively correlated with *ACE2* expression in two independent cohorts. The interferon-associated down-regulation of *ACE2* was also identified in lung tissue in a murine model of SARS-CoV-1 infection. Performing a genome-wide eQTL analysis for *ACE2* expression in the virus infected liver biposies, we found that *ACE2* expression is associated with host genetic variation in the *IFNL* region on chromosome 19q13.2 which has previously been associated with differential hepatic ISG expression in chronic HCV infection. We also observed that increase in age and presence of liver cirrhosis is significantly associated with increase in *ACE2* expression. Due to the conserved pattern of down-regulation of *ACE2* in presence of up-regulation of ISGs across tissues, inflammatory responses, infections and species, we conclude that *ACE2* is a negatively-regulated ISG and the genetic variation in the *IFNL* locus which modulates ISGs expression in viral infection may potentially play a role in SARS-CoV-2 pathogenesis.

## Results

To detect common regulatory mechanisms, biological function and the context of *ACE2* expression, we performed correlation analysis accounting for multiple testing to identify genes correlated with *ACE2* expression in virus-infected livers. We observed large correlation coefficients (maximum of 0.6 and minimum of -0.5) and detected 1530 genes significantly correlated with *ACE2* expression at 1% false discovery rate (FDR) and with correlation coefficients of > 0.3 or < -0.3. Considering separately the genes that were positively correlated (N=1362) and those that were negatively correlated (N=168) with *ACE2* expression (**Supplementary Tables 1** and **2**), we performed a gene set enrichment analysis, observing that genes involved in type I interferon signalling pathways were enriched among genes negatively correlated with *ACE2* expression (**Figures 1a** and **1b, Supplementary Table 3**). We also observed that genes involved in extracellular structure organisation were enriched among genes positively correlated with *ACE2* expression (**Supplementary Figure 1** and **Supplementary Table 4**). We used an independent data set of liver biopsies^9^ from 28 patients (6 non-HCV infected controls and 22 HCV infected cases, GSE84346) and replicated concordant correlation signs for 162 of the 168 genes negatively correlated with *ACE2* (**Figure 1c** and **Supplementary Table 5**). This represents a significant enrichment of concordant correlation signs relative to the null hypothesis of no association between correlation coefficients in the two datasets (p=2.2×10^-16^, binomial test). In this replication cohort 18 patients had two biopsies taken, one before and another after treatment with pegylated IFN-α at different time points (N_4 hours_=4, N_16 hours_=3, N_48 hours_=3, N_96 hours_=3, N_144 hours_=5). At all time points we observed a down-regulation of *ACE2* expression (**Figure 1d**), with the biggest median drop at 16 hours post injection. These reductions were nominally significant at 48 and 144 hours after IFN-*α* treatment (paired t-test, **Figure 1e**), but across all time points represent a highly significant down-regulation of *ACE2* after IFN-*α* treatment (p=6.5×10^-5^, paired t-test).

**Figure 1:**
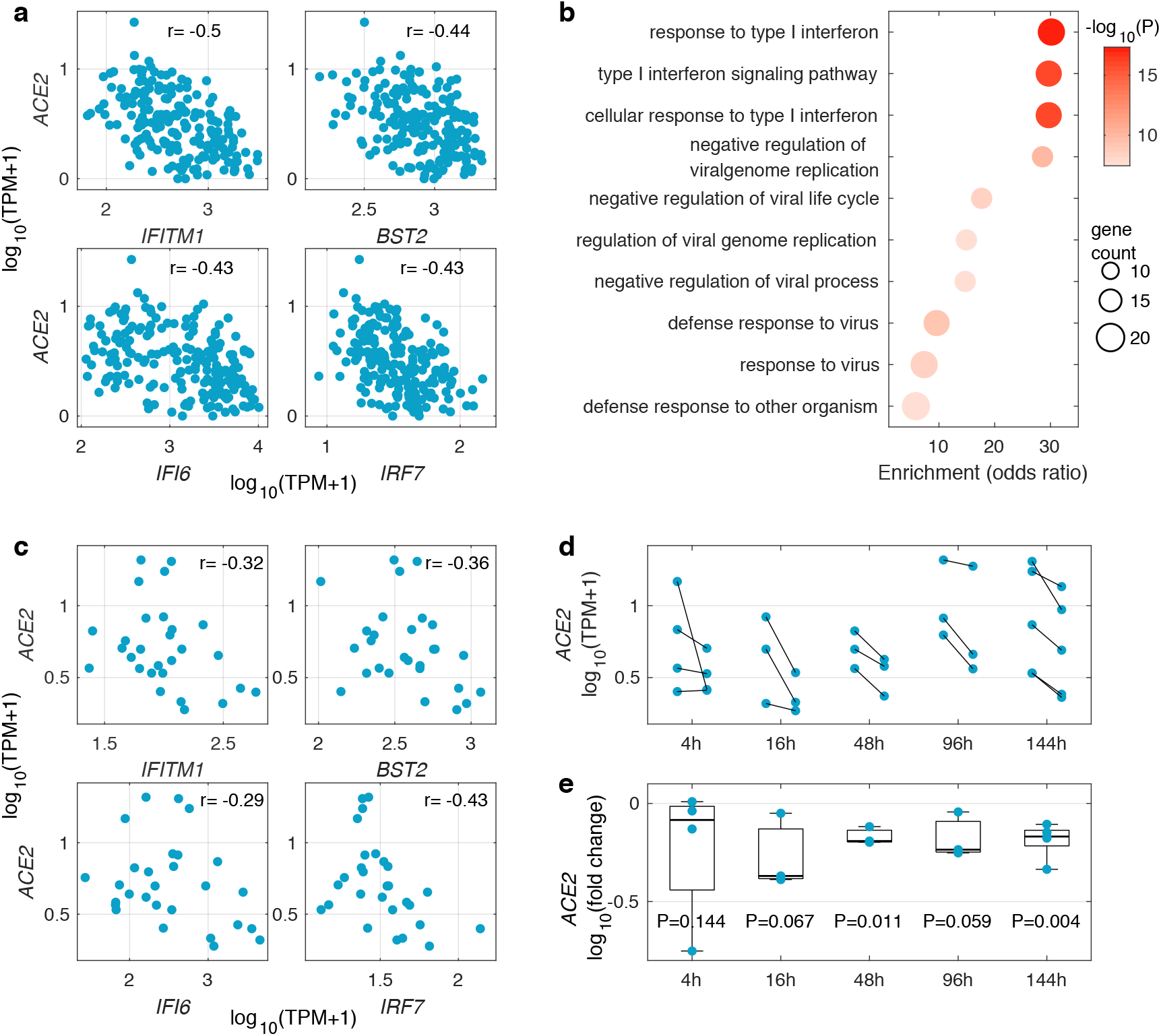
Negative correlation of transcript levels of *ACE2* with interferon-stimulated genes in HCV infected liver biopsies. (**a**) Expression of four representative interferon-stimulated genes (ISG) and their observed negative correlation with *ACE2* expression. Pearson’s correlation coefficient is shown for each gene. (**b**) Gene Ontology (GO) gene set enrichment analysis among genes with significant negative correlation with *ACE2* expression (at false discovery rate of 1% and correlation coefficient of < -0.3). Only the top ten enriched gene sets are shown (from a total of 22 gene sets), which are all ISGs related pathways. (**c**) Expression of the four representative ISGs and their observed negative correlation with *ACE2* in an independent HCV infected liver biopsy data set. Pearson’s correlation coefficient is shown for each gene. (**d**) *ACE2* expression in paired liver biopsy samples from before and after pegylated IFN*-α* treatment taken at different time points post treatment. (**e**) Log fold change (log_10_ (post-treatment / pretreatment)) of *ACE2* expression after pegylated IFN-*α* injection. At all time points we observe a reduction in mean *ACE2* expression with the biggest median drop at 16 hours. The reported p-values are from paired t-tests for each time point. Combining all time points, down-regulation of *ACE2* after IFN-*α* treatment is highly significant (p=6.5×10^-5^, paired t-test).

To further explore the negative correlation of ISGs with *ACE2* expression in a known site of SARS-CoV-2 replication, we explored the relationship between *ACE2* and ISGs expression in the gastrointestinal (GI) tract in a gene expression study of terminal ileum biopsies in inflammatory bowel disease (IBD) in treatment-naive young donors (RISK cohort^14^, GSE57945). In intestinal biopsies, there was a striking decrease of *ACE2* expression with increasing severity of inflammation that was independent of the abundance of transcriptional markers of epithelial identity^15^ (**Figure 2a** and **Supplementary Figure 2a**) and ISGs had increasing expression with rise in disease activity and were negatively correlated with *ACE2* expression (**Figures 2b, 2c** and **Supplementary Figure 2b**). Genes associated with epithelial cell structure and function were enriched among genes that were positively correlated with *ACE2* in both liver and intestine, while genes associated with type I interferon signalling pathways were enriched among genes that negatively correlated with *ACE2* expression in both tissues (**Supplementary Figure 3**). These data were supported by analysis of a second independent IBD cohort^16^ (GSE137344, **Supplementary Figure 4**).

**Figure 2:**
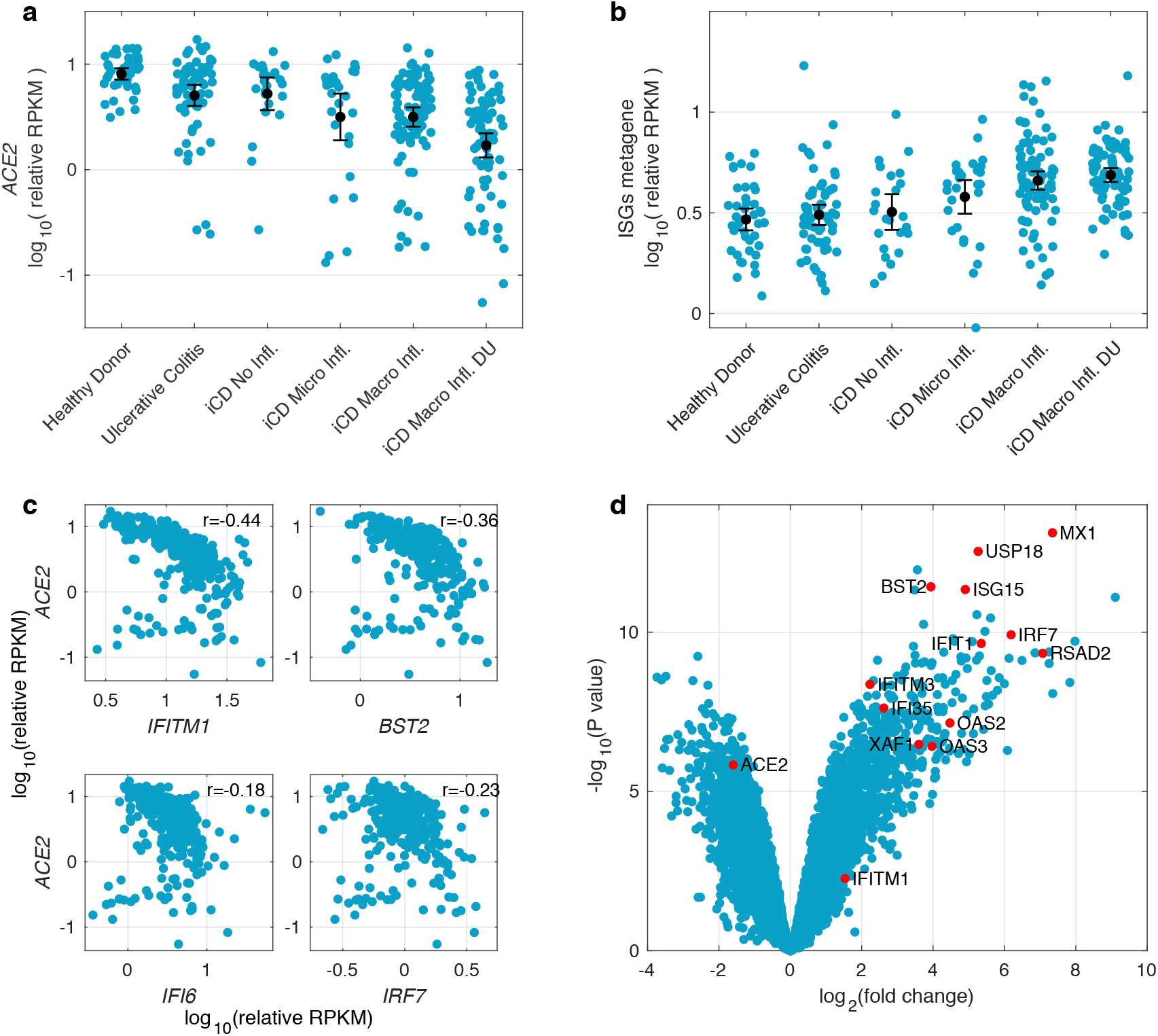
Conservation of negative correlation of *ACE2* expression with ISGs across tissues, conditions and species. (**a**) *ACE2* expression in terminal ileum biopsies of the RISK cohort grouped based on disease severity and histologic assessment of inflammation. The black dots and lines indicate the mean and its 95% confidence interval in each group (Ulcerative Colitis without ileal involvement; iCD = ileal Crohn’s Disease; Micro Infl. = Microscopic Inflammation, Macro Infl. = Macroscopic Inflammation; DU = Deep Ulcers). Data are shown as RPKM relative to the epithelial cell identity metagene (see methods) (**b**) Expression of genes which are part of the Gene Ontology category “response to type I interferon” (GO ID: 0034340) and which in the liver are negatively correlated with *ACE2* (FDR of 1% and correlation coefficient of < -0.3) in the RISK cohort stratified by disease severity. The black dots and lines show the mean and its 95% confidence interval for each group. (**c**) Negative correlation of representative ISGs with *ACE2* in the RISK cohort. Pearson’s correlation coefficients are shown. (**d**) Volcano plot of lung tissue differential gene expression pattern induced by SARS-CoV-1 infection in mouse model vs. mock. *ACE2* and representative ISGs which are part of the Gene Ontology category “response to type I interferon” (GO ID: 0034340) and which in the liver are negatively correlated with *ACE2* are indicated by red dots.

Since the pattern of gene expression incorporating down-regulation of *ACE2* in presence of ISGs was consistent in two models of viral chronic infection and/or inflammation in different tissue, we addressed whether a similar pattern of gene regulation was observed in lung tissue using data from mouse models of SARS-CoV-1 infection^17^ (GSE59185). Indeed we observed in SARS-CoV-1 infected lung the same associated down-regulation of *ACE2* in the presence of up-regulation of classical ISGs (**Figure 2d**).

In the context of chronic HCV infection, ISG induction varies considerably between individuals with some patients showing constant ISGs expression at high levels while others show almost no detectable induction of innate immune system^7,8^. This differential expression of the ISGs is strongly associated with genetic variants in the *IFNL* locus on chromosome 19q13.2. This locus has also been strongly associated with spontaneous clearance of HCV in acute phase of infection^18^, IFN-*α* treatment response in chronic phase of infection^13^, viral load and viral evolution^19,20^. The causal variant is likely to be the dinucleotide exonic variant rs368234815 in *IFNL4*^21^. This variant [∆G > TT] results in a frameshift, abrogating production of functional IFN-λ4 protein. Lack of production of IFN-λ4 (carrying rs368234815 TT/TT genotype) is associated with no or low levels of expression of liver ISGs and higher viral load and paradoxically with higher rates of spontaneous clearance and treatment response to IFN-*α* and direct acting antivirals^21,22^. Lack of production of IFN-λ4 is also associated with better outcome of RNA virus respiratory tract infections (including coronaviruses) in children^23^. It has been shown that IFN-λ4 is highly conserved in mammals and therefore functionally relevant, but in humans, the pseudogene (rs368234815 TT allele) has a gradient in frequency that rises from Africa (0.29–0.44) to Europe (0.58–0.77) and reaches near fixation in East Asia (0.94–0.97) indicating positive selection has favoured the elimination of IFN-λ4 in humans^24^. The molecular link between the genotype and the phenotype remains to be discovered.

To understand the link between host genetics and *ACE2* expression, we used genotyped autosomal SNPs in the host genome to undertake a genome-wide eQTL analysis for the expression of *ACE2* in virus-infected liver. Due to a dominant effect of *IFNL4* locus, we used both additive and dominant genetic models using linear regression and adjusted for population structure by including the first five host genetic principal components (PCs) as covariates. We also added age, sex and liver cirrhosis status as covariates to account for possible confounding. There was no inflation in the association test statistics (**Supplementary Figure 5**). We used a threshold of 5×10^-8^ to decide on significance. Across the human genome, in both dominant and additive analysis, the most associated signals were observed for three SNPs in the *IFNL* locus (**Figure 3a**, **Supplementary Figures 6, 7, 8** and **Supplementary Table 6**). For all three SNP the dominant model had lower p-values (rs12980275, P_dom_=9.9×10^-11^, P_add_=1.6×10^-8^; rs8103142, P_dom_=7.8×10^-10^, P_add_=2.5×10^-8^; rs12979860, P_dom_=3.9×10^-9^, P_add_=6.5×10^-8^). These SNPs are in high linkage with each other and with the rs368234815 SNP (not typed in our genotyping array) in European populations (**Supplementary Figure 9**).

**Figure 3:**
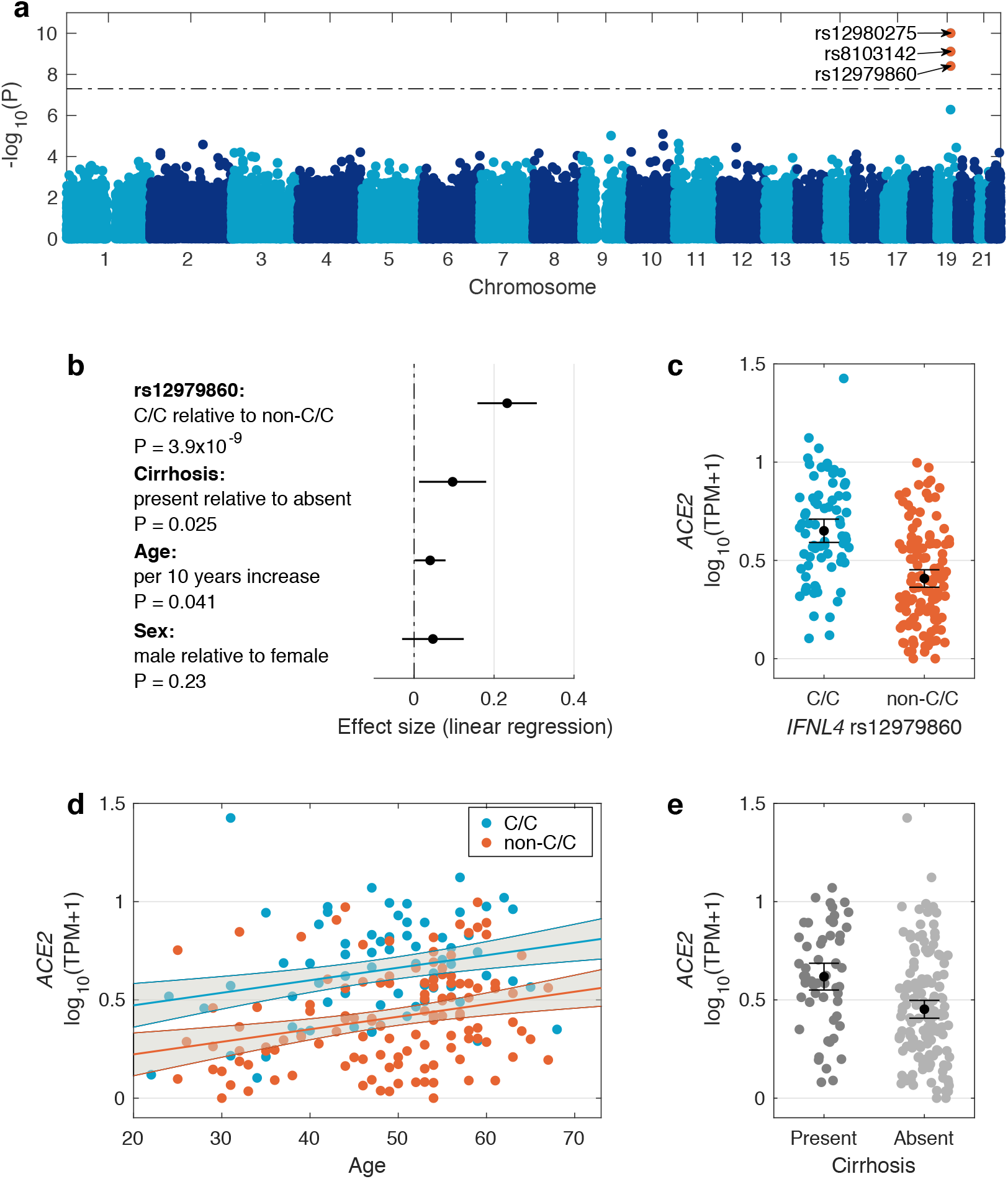
Impact of host genetics and other factors on *ACE2* expression in HCV infected liver. (**a**) Manhattan plot of association between host genetic variation and *ACE2* expression in virus infected liver biopsies using a dominant genetic model. The dashed line indicates 5×10^-8^ significance threshold. Significant SNPs are coloured red and their ID is shown. (**b**) Forest plot of the effect sizes of SNP rs12979860 (dominant model C/C vs. C/T and T/T genotypes), cirrhosis status, age and sex on *ACE2* expression. The black circles indicate the point estimate of the effect sizes and the black lines indicate their 95% confidence interval. (**c**) Distribution of *ACE2* expression stratified by SNP rs12979860 genotypes (dominant model). Black circle shows the mean and the lines indicate its 95% confidence interval. (**d**) The relationship between *ACE2* expression and age. The blue and red lines show the linear regression fit (for C/C and non-C/C genotypes respectively) and the grey area indicates their 95% confidence interval. (**e**) Distribution of *ACE2* expression stratified by host liver cirrhosis status. The black dot and lines indicate the mean and its 95% confidence interval.

To further understand the impact of polymorphisms in the host *IFNL4* gene and other host factors on *ACE2* expression in presence of viral infection, we focused on the impact of *IFNL4* SNP rs12979860 on *ACE2* expression (**Figure 3b**). This SNP is an *IFNL4* intronic SNP and closest to rs368234815 SNP among the three associated SNPs, where rs12979860 C allele is in linkage with rs368234815 TT allele. Using the dominant genetic model: C/C vs. C/T and T/T genotypes (i.e. those that do not produce IFN-λ4 protein [associated with low or no hepatic ISG expression] and those that do produce IFN-λ4 protein [associated with high hepatic ISG expression]), we observed significantly higher expression of *ACE2* (P=3.9×10^-9^) in individuals with C/C vs. non-C/C genotypes (Figure 3c). Additionally, we observed that *ACE2* expression increased with age (P=0.04) in both C/C and non-C/C patients (Figure 3d) and patients with cirrhosis on average had higher *ACE2* expression (P=0.02, Figure 3e).

We also investigated the impact of the *IFNL4* SNP rs12979860 genotypes on *TMPRSS2, CTSB* and *CTSL* genes which may also be needed for viral entry^1^. Linear regression using a dominant genetic model (C/C vs. C/T and T/T) was used for association testing and the first five host PCs as well as cirrhosis status, age and sex were added as covariates to the analysis. We observed that these three genes had much higher expression levels in the liver relative to *ACE2*, however their expression was not significantly associated with SNP rs12979860 genotypes (**Supplementary Figure 10**). Both *CTSB* and *CTSL* genes had significantly lower expression in patients with cirrhosis (P_CTSB_=0.02, P_CTSL_=7×10^-3^), while *TMPRSS2* had higher level of expression in cirrhotic patients (P=3×10^-3^, **Supplementary Figure 11**).

## Discussion

To understand the heterogeneity of *ACE2* expression *in vivo* and detect common regulatory mechanisms in presence of RNA virus infection, we measured the correlation between *ACE2* expression and nearly 15000 other genes expressed in HCV infected liver biopsies. We observed a down-regulation of *ACE2* in the presence of up-regulation of classical ISGs. We also observed down-regulation of *ACE2* in liver biopsies after pegylated-IFN-alpha injection with the biggest drop at 16 hours post injection. The same observation was confirmed in the gastrointestinal tract in an inflammatory condition and in lung in a murine model of SARS-CoV-1 infection. This indicates a robust maintenance of the transcriptional down-regulation of *ACE2* in presence of ISGs up-regulation across infections, inflammatory conditions, tissues and species.

In the context of chronic HCV infection, hepatic ISG expression varies among individuals with some patients having no or low interferon response while others having full activation of ISGs. This differential innate immune activation is highly associated with the genetic variation in the *IFNL* locus^7,8^. To understand the impact of this locus and other host genetic factors on *ACE2* expression in the presence of RNA virus infection, we performed a genome-wide eQTL analysis for *ACE2* expression in our liver biopsies. Using infected tissue is important, since genetically driven differences in innate immune responses are only likely to be observed when innate immune responses are triggered. We observed that genome-wide, host genetic polymorphisms in the *IFNL* region were significantly associated with *ACE2* expression in the presence of viral infection. The impact of this locus is likely to be mediated through the differential ISGs activation, associated with this locus.

The likely causal mechanism is the variant rs368234815 [∆G > TT], which results in a frameshift and abrogates production of IFN-λ4 ^21^. In the context of HCV infection, production of IFN-λ4 is associated with high hepatic ISG expression (low *ACE2* expression) and low viral load, but paradoxically with lower rates of spontaneous clearance in acute phase of infection and lower rates of response to treatment in the chronic phase of infection. The functional link between the locus and the phenotypes has not been fully defined, in part due to limitations in detection of IFNL proteins and mRNAs in the liver biopsies of patients with chronic HCV infection ^25,26^.

Interferon lambda receptor (*IFNLR1*) is largely restricted to tissues of epithelial origin^27,28^, therefore, IFN-λ proteins (type III IFN) may have evolved specifically to protect the epithelium. Overall, *INFL* genes lead to a pattern of gene expression which is similar to type I interferon genes, but the time course and pattern of expression may vary^10^. This has been explored in HCV, where a slower, but sustained impact of *IFNL* signalling is seen^29^. *In vitro* studies have revealed that ISG expression and anti-viral activity induced by recombinant *IFNL4* are comparable to that induced by *IFNL3^30^*, although the tight regulation of *IFNL4*^31^ may impact on its ability to induce a rapid antiviral state^32, 22^. However, once established, the *IFNL4* transcriptional module may also be highly sustained (as seen here and in other HCV cohorts^33^) and also noted elsewhere, e.g. after childbirth^34^.

In mice, the type III IFN response is restricted largely to mucosal epithelial tissues, with the lung epithelium responding to both type I and III IFNs^35^ and intestinal epithelial cells responding exclusively to type III IFNs. Among nonhematopoietic cells, epithelial cells are potent producers of type III IFNs. In mouse models, type III IFNs seem to be the primary type of IFN found in the bronchoalveolar lavage in response to influenza A virus infection and play a critical role in host defence^36^. The data from the GI tract indicate that this gene expression pattern is conserved amongst tissues, consistent with emerging data^37^.

We have presented evidence that *ACE2* may be negatively regulated by IFNs *in vivo*. We have also demonstrated that *ACE2* expression in presence of RNA virus infection is modulated by genetic variation in the *IFNL* region. This regulation is likely due to confirmed differential activation of innate immune system in response to RNA virus infection associated with this region. This region is also associated with outcome of RNA virus respiratory tract infection in children^23^. Therefore given the prominent role of type III interferons in defence of epithelial surfaces such as that in the lung from viral infections, we hypothesise that the genetic variation in the *IFNL* region to also play an important role in modulating innate immune responses to SARS-CoV-2 infection. High ISGs level and down-regulation of *ACE2* may limit the ability of SARS-CoV-2 and other related coronaviruses to enter cells, but may, if sustained, also have impacts on inflammation. Indeed *ACE2^-/-^* mice suffer from enhanced disease following virus infection of the lung through an angiotensin-driven mechanism^38^.

These data are derived from an *in vivo* assessment and the down-regulation of *ACE2* is consistent across tissues and species. The data are also potentially consistent with up-regulation of *ACE2* seen in early time points by IFN-*α in vitro*^39^. Likely the explanation for the difference is that the regulation of this physiologic receptor in an *in vivo* setting is distinct from studies *in vitro* - but the full kinetics of this need further study during natural infection.

This study is relevant to the expression of *ACE2* during SARS-CoV-2 infection. Although we did not study this directly in the respiratory tract, such studies should be performed to confirm these data. Furthermore the overall impact of *IFNL4* polymorphism on the clinical course should be assessed, especially given the very variable distribution of *IFNL4* alleles in different ethnic groups^13,24^. Finally, the genetic data add weight to the idea of a careful exploration of IFN-λ pathways in therapy for SARS-CoV-2^40^.

## Data Availability

The liver gene expression read counts are submitted to Gene Expression Omnibus (GEO) under accession number GSE149601. The raw FASTQ files are currently being deposited in the European Genome-phenome Archive and the accession number will be provided when the submission is finalised. Human genotype data underlying this manuscript are deposited in the European Genome-phenome Archive under accession code EGAS00001002324. Due to patient privacy concerns the gene expression FASTQ files and the human genotype data can only be accessed by making an application to the data access committee. The Information on access to the study data is available at http://www.stop-hcv.ox.ac.uk/data-access.

## Acknowledgements

The authors would like to thank Gilead Sciences for the provision of samples and data from the BOSON clinical study for use in these analyses. The authors would also like to thank HCV Research UK (funded by the Medical Research Foundation) for their assistance in handling and coordinating the release of samples for these analyses. The authors would also like to thank ISARIC-4C investigators for their discussion and contributions to this study. This work was funded by a grant from the Medical Research Council (MR/K01532X/1 – STOP-HCV Consortium). The work was supported by Core funding to the Wellcome Centre for Human Genetics provided by the Wellcome Trust (203141/Z/16/Z). This work was also supported by the Medical Research Council [grant number MC_PC_19059] and a strategic award from the Wellcome Trust (211276/Z/18/Z – WSSS). MAA is supported by a Wellcome Trust Sir Henry Dale Fellowship (220171/Z/20/Z). PK is supported as a Wellcome Trust Senior Investigator (WT 109965MA) and an NIHR Senior Investigator. PK is affiliated to the National Institute for Health Research Health Protection Research Unit (NIHR HPRU) in Emerging and Zoonotic Infections at University of Liverpool in partnership with Public Health England (PHE), in collaboration with Liverpool School of Tropical Medicine and the University of Oxford. EB was funded by the Medical Research Council UK, the Oxford NIHR Biomedical Research Centre and is an NIHR Senior Investigator. The work was also supported by the NIHR Biomedical Research Centre, Oxford. The views expressed are those of the author(s) and not necessarily those of the NHS, the NIHR, the Department of Health or Public Health England. The authors thank Jan Rehwinkel for his contributions to the manuscript preparation and David Klenerman (University of Cambridge) for his contribution to the high throughput sequencing platform that underpins this work.

## Code availability

The R and MATLAB code used to generate the results and figures from the primary analyses described below are available from the authors on request.

## Methods

### Boson patient cohort

For this study, we used patient data from the BOSON cohort that has been described elsewhere^41^. All patients provided written informed consent before undertaking any study-related procedures. The BOSON study protocol was approved by each institution’s review board or ethics committee before study initiation. The study was conducted in accordance with the International Conference on Harmonisation Good Clinical Practice Guidelines and the Declaration of Helsinki (clinical trial registration number: NCT01962441).

### RNA extraction, library prep, sequencing and mapping for the BOSON cohort

Liver biopsy samples were available for 198 patients. Total RNA was extracted from patient liver biopsies at baseline (pre-treatment) using RNeasy mini kits (Qiagen). Briefly, liver biopsy samples were mechanically disrupted in the presence of lysis buffer and homogenized using a QIAshredder. Tissue lysates were then centrifuged and clarified supernatants were transferred into new microcentrifuge tubes (pellets were discarded). Next, 1 volume of 70% ethanol was added to the lysates and samples were mixed by gentle vortexing. 700uL of sample was then transferred into RNeasy spin columns (with 2mL collection tubes) and centrifuged at 10000 rpm for 15 seconds. Column flow-through was discarded. DNase digestion was subsequently performed to eliminate any contamination from genomic DNA. 80uL of DNase I solution (10uL DNase I stock + 70uL Buffer RDD) was added directly to RNeasy spin columns and incubated at room temperature for 15 minutes. Following DNase incubation, the columns were washed with 350uL of Buffer RW1 and centrifuged at 10000 rpm for 15 seconds. Flow-through was discarded and 500uL of Buffer RPE was added to the spin columns. Columns were then centrifuged again at 10000 rpm for 15 seconds and flow-through was discarded. An additional 500uL of Buffer RPE was added to the spin columns and columns were centrifuged at 10000 rpm for 2 minutes. Finally, spin columns were transferred into new microcentrifuge tubes and 30uL of RNase-free water was added directly to the column membrane. Columns were then centrifuged at 10000 rpm for 1 minute to elute the RNA.

RNA yield was quantified using a NanoDrop spectrophotometer. Selected samples were also run on an Agilent TapeStation system to assess RNA quality and purity. Library preparation from purified RNA samples was performed using the Smart-Seq2 protocol^42^, used along with previously described indexing primers during amplification^43^.

High-throughput RNA sequencing of prepared libraries was performed on the Illumina HiSeq 4000 platform to 75bp PE at the Wellcome Center for Human Genetics (Oxford, UK). Reads were trimmed for Nextera, Smart-seq2 and Illumina adapter sequences using skewer-v0.1.125^44^. Trimmed read pairs were mapped to human genome GRCh37 using HISAT2 version 2.0.0-beta^45^. Uniquely mapped read pairs were counted using featureCounts^46^, subread-1.5.0^47^, using exons annotated in ENSEMBL annotations, release 75. Mapping QC metrics were obtained using picard-tools-1.92 CollectRnaSeqMetrics.jar. Three samples were excluded after QC checks due to low sequencing depth which left 195 samples for analysis. Genes were filtered using the criteria of having a count per million (CPM) of 1.25 in at least 10 samples to remove low expressed genes. Function cpm from edgeR^48^ version 3.20.9 was used to calculate the CPM values. After removing low expressed genes we were left with 14882 genes. To normalise for library size and gene length, transcripts per million (TPM) values were calculated from unique mapped read counts and log_10_ (TPM+1) was used in the analysis.

For the replication cohort^9^ (GSE84346), the read counts were downloaded from GEO. Gene expression data for 46 liver biopsy samples from 28 individuals were available for this data set. 22 individuals had chronic HCV infection while 6 individuals did not have HCV infection and were enrolled as controls. Among the 22 individuals with chronic HCV infection, 18 individuals were treated with pegylated interferon-alpha and a second biopsy was taken post-treatment. Genes were filtered using the criteria of having a count per million (CPM) of 1.25 in at least 10 pre-treatment samples to remove low expressed genes. After removing low expressed genes we were left with 14661 genes. To normalise for library size and gene length, transcripts per million (TPM) values were calculated from unique mapped read counts and log_10_ (TPM+1) was used in the analysis.

### Host genotyping

Host genome-wide genotyping was performed on 567 patients from the BOSON cohort as described previously^20^. Briefly, genomic DNA was extracted from buffy coat using Maxwell RSC Buffy Coat DNA Kit (Promega) as per the manufacturer’s protocol and quantified using Qubit (Thermofisher). DNA samples from patients were genotyped using the Affymetrix UK Biobank array^20^. Both liver RNA trascriptomic and human genome-wide SNP data were obtained on a total of 190 patients of mainly White self-reported ancestry infected with HCV subtype 3a. After quality control and filtering of the human genotype data, approximately 330,000 common SNPs with minor allele frequency greater than 5% were available for analysis.

### Statistical analysis

For the BOSON Cohort, Log_10_(TPM+1) values were calculated and used to estimate Pearson’s correlation coefficient between *ACE2* and all other genes. The qvalue package in R was used to calculate false discovery rate. We used FDR of 1% and correlation coefficient of >0.3 or <-0.3 to decide on genes significantly correlated with *ACE2*. To test for enrichment we used enrichGO function from the clusterProfiler package^49^, limiting the analysis to GO “biological process” class and maximum gene set size of 500.

An independent liver biopsy dataset (GSE84346) was used to replicate the negative correlation of *ACE2* with 168 genes found in the BOSON cohort. We used the Log_10_ (TPM+1) values from the 28 baseline liver biopsy samples and calculated Pearson’s correlation coefficient between *ACE2* and the 168 genes. Assuming a null hypothesis of no association between correlation coefficients signs in the two data sets, one can use a binomial test to assess this null hypothesis where the number of trials (n) is 168 and the probability of a negative correlation sign was estimated from the BOSON cohort by dividing the number of genes with negative correlation with *ACE2* divided by total number of genes (14882). To test the hypothesis of down-regulation of hepatic *ACE2* expression when treated with pegylated interferon-alpha, we used the 18 individuals with liver biopsy samples taken before and after the treatment. We used one-sided paired t-test to perform hypothesis testing for each of the time points and across all time points.

To test for association between autosomal human SNPs and *ACE2* expression (Log_10_ (TPM+1)), we performed linear regression using PLINK^50^ version 1.9 using additive and dominant genetic models adjusted for the human population structure by adding the first five genetic principal components as covariates. We also added host cirrhosis status, age and sex as covariates to the analysis. To assess the impact of age per 10 years increase, we divided the age by 10 before adding it as a covariate. For 190 patients both host genome-wide genotyping data and hepatic *ACE2* expression data were available. We used a significance threshold of 5×10^-8^.

To test for association between *IFNL4* SNP rs12979860 genotypes and expression of *CTSL, CTSB* and *TMPRSS2* genes, we used linear regression and added the first five host genetic principal components as covariate to account for population structure. Patient cirrhosis status, age and sex were also addes as covariates in this analysis.

### RISK cohort

The RISK study is an observational prospective cohort study with the aim to identify risk factors that predict complicated course in pediatric patients with Crohn’s disease as previously described^51^. The RISK study recruited treatment-naive patients with a suspected diagnosis of Crohn’s disease. The Paris modification of the Montreal classification were used to classify patients according to disease behaviour (non-complicated B1 disease (non-stricturing, non-penetrating disease); complicated disease, composed of B2 (stricturing) and/or B3 (penetrating) behaviour) as well as disease location (L1, ileal only, L2, colonic only, L3, ileocolonic and L4, upper gastrointestinal tract). 322 samples were investigated with ileal RNA-seq. Individuals without ileal inflammation were classified as non-IBD controls. Patients with Crohn’s disease were followed over a period of 3 years. Patients were largely of European (85.7%) and African (4.1%) ancestry. RPKM expression values for the RISK cohort^51^ were retrieved from GEO (GSE57945). The dataset was filtered to (n=19,556) genes that had an expression value ≥ 0.1 in >10% of the patients.

### Statistical analysis

To account for the potential loss of epithelial cells contribution to gene expression a metagene score was generated based on the average expression of epithelial identity genes^15^. RPKM data were transformed and presented as: RPKM+1/epithelial cell metagene. For the ISG metagene score, we used 17 genes which were significantly negatively correlated with *ACE2* in the liver and were part of the GO term “response to type I interferon” (GO ID: 0034340). For the intersection of *ACE2* correlated gene expression, genes were ranked based on their Pearson’s correlation coefficient with *ACE2* for each patient subgroup. Intersected lists of *ACE2* expression positively (Pearson’s correlation coefficient > 0.5) and negatively (Pearson’s correlation coefficient < -0.5) correlated genes were extracted (positive correlation: n = 2067; negative correlation: n = 2264). BOSON liver *ACE2* expression and RISK *ACE2* expression positively and negatively correlated gene sets were intersected based on Entrez gene identifiers using Cytoscape (version 3.7.1) and visualized using the Cytoscape Venn and Euler Diagrams (Version 1.0.3) plugin (http://apps.cytoscape.org/apps/vennandeulerdiagrams). Functionally grouped networks of terms and pathways were analysed using the Cytoscape (version 3.7.1) ClueGO (version 2.5.6) and CluePedia (version 1.5.6) plug-in^52^. The analysis was performed by accessing the Gene Ontology Annotation (GOA) Database for Biologic processes, Cellular components, Immune system processes and Molecular function, the Reactome pathways database (https://reactome.org/) and the KEGG database (https://www.genome.jp/kegg/pathway.html). Only pathways with an adjusted enrichment p-value ≤ 0.05 were considered (Two-sided hypergeometric test, Bonferroni step down p-value correction). GO terms were grouped based on the highest significance when more than 50% of genes or terms were shared. The filtered RISK gene expression data (n=19,556; expression value ≥ 0.1 in >10% of the patients) served as reference gene set.

Resources for statistical analysis and data visualization:

Prism version 8.0 (GraphPad Software)

Excel for Mac Version 15.32 (Microsoft)

Cytoscape 3.7.1 (https://cytoscape.org/)

Cytoscape 3.7.1 plugin ClueGO (Version 2.5.6)

Cytoscape 3.7.1 plugin CluePedia (version 1.5.6)

Cytoscape 3.7.1 plugin Venn and Eluler Digrams (Version 1.0.3)

Morpheus (https://software.broadinstitute.org/morpheus/)

R (Version 3.6.1)

RStudio (Version 1.2.5001)

Specific statistical tests applied in this study are described in the respective figure legends. The level of statistically significant difference was defined as p ≤ 0.05.

### GENESIS cohort

GENESIS is funded by the National Institute of Diabetes and Digestive and Kidney Diseases and managed by Emory University for the recruitment of self-identified African American subjects with IBD^53^. We used a subset of 195 GENESIS cohort subjects with ileal transcriptomic profiles as an additional replication cohort to test for negative correlation of ACE2 expression with interferon-stimulated genes expression. Pearson correlation tests between normalized expression values for *ACE2* and four ISGs confirmed that this pattern of negative correlation is also observable in a cohort enriched for African American ancestry. This dataset includes 158 IBD patients along with 37 controls. Subjects with ileal inflammation were included as IBD, while non-IBD controls did not have ileal inflammation. This dataset is enriched for African American ancestry (70%), and gender was equally distributed. Full descriptions of age, gender, race, disease status and other phenotypic information are available in a prior publication^16^. Additionally, ileal transcriptomic profiles sequenced on the NextSeq 550 platform are available in the GEO repository (GSE57945) for all subjects.

### Data analysis for SARS-CoV-1 mouse model

The mouse lung tissue microarray data were downloaded from GEO using the accession number GSE59185^17^. In case of multiple probes per gene, they were collapsed into a single feature, which resulted in 21217 features. Three lung tissue samples were from mice infected with wild type virus and three lung tissue samples were from mock infection. We then used LIMMA^54^ to perform differential gene expression analysis between these two conditions. The log fold change and the p-values were used to make a volcano plot. The genes highlighted as red on the volcano plot are 17 genes which were significantly negatively correlated with *ACE2* in the human liver and were part of the GO term “response to type I interferon” (GO ID: 0034340).

## Notes

### Competing Interest Statement

The authors have declared no competing interest.

### Author Declarations

The BOSON study protocol was approved by each institution review board or ethics committee before study initiation. The study was conducted in accordance with the International Conference on Harmonisation Good Clinical Practice Guidelines and the Declaration of Helsinki (clinical trial registration number: NCT01962441).

